# Cerebrospinal fluid cellular dynamics following modified interval dosing natalizumab therapy in multiple sclerosis

**DOI:** 10.64898/2026.11.25.26368928

**Authors:** Masashi Fujita, Tina Roostaei, Claudiu Diaconu, Marie Deffner, Ya Zhang, Ilya Kister, Asya I. Wallach, Kaho Onomichi, Juliana Oyegunle, Fatoumata Diallo, YunDuo Liu, Ellen Cahir-McFarland, Tamar Kalina, Vilas Menon, Regina Berkovich, Hanane Touil, Gerd Meyer zu Hörste, Heinz Wiendl, Susan Goelz, Lana Zhovtis Ryerson, Philip L. De Jager, Nicholas Schwab, Claire S. Riley

## Abstract

**Background:** Natalizumab, an α4-integrin monoclonal antibody, a highly effective relapsing multiple sclerosis (MS) therapy, carries a small but clinically important risk of progressive multifocal leukoencephalopathy (PML). Previous studies have suggested that alternative dosing paradigms can reduce PML risk while maintaining therapeutic efficacy.

**Objectives:** Clarify how immune cell dynamics changes with the alternative dosing.

**Methods:** We investigated alternative dosing paradigms by profiling cerebrospinal fluid (CSF) immune cell dynamics across: (1) standard interval dosing (SID, every 4 weeks), (2) extended interval dosing (EID, every 6 or 8 weeks), and (3) annual 12-week gap (aGAP). Using single-cell RNA sequencing, we analyzed 46 CSF samples from participants across dosing regimens, untreated MS controls, and one non-natalizumab-related PML case.

**Results:** Compared with controls, natalizumab reduced proportions of Treg, memory T cells, B cells, and plasmacytoid dendritic cells, while increasing tissue-resident memory T and microglia-like myeloid cells. EID and aGAP were associated with higher proportions of peripheral immune populations, including CD4+ T cells with interferon response signatures (CD4+IFN+), potentially enhancing antiviral immunity. The PML case exhibited elevated CD8+ central memory T and CD56^bright^ NK cells proportions in the CSF, suggesting their role in viral control.

**Conclusions:** These findings support that EID and aGAP allow immune reconstitution.

## Introduction

Natalizumab is a humanized monoclonal antibody directed against very late antigen 4 (VLA-4) that blocks the interaction of α4β1 integrin on circulating leukocytes with vascular cell adhesion molecule 1 (VCAM-1) on the vascular endothelium, thereby preventing leukocyte trafficking across the blood brain barrier. This mechanism of preventing leukocyte egress into the central nervous system (CNS) has proven highly effective for preventing relapses in multiple sclerosis (MS)(1). The emergence of the potentially lethal viral infection progressive multifocal leukoencephalopathy (PML) as a serious adverse event prompted transient withdrawal from the US market in 2005. Natalizumab returned to use in 2006 with a robust risk evaluation and mitigation strategy in place.

PML results from an opportunistic infection of the CNS by the John Cunningham virus (JCV) in individuals undergoing natalizumab treatment and individuals with a dysfunctional immune system such as inflammatory diseases or native immunodeficiencies (2, 3). It was recognized most often in profound CD4+ T cell depletion from HIV/AIDS (4).

Reduced CNS infiltration of lymphocytes by natalizumab retains activated T cells in the periphery (5), but it also impairs immune surveillance for CNS infections. In the case of natalizumab, this perturbation appears relatively specific, affecting the risk of PML but not driving a significant increase in other opportunistic infections of the CNS. Further, while approximately 50-60% of individuals with MS have antibodies against JCV, the rate of PML remains relatively low (6, 7). However, natalizumab has a clinically meaningful impact on PML risk because treatment of natalizumab-associated cases of PML could result in rapid re-entry of lymphocytes that precipitates severe potential sequelae including death (8).

Refined titration of natalizumab could reduce the risk of PML further while preventing relapses of MS. A randomized controlled trial of standard and extended interval dosing (SID, EID) as well as observational studies of natalizumab dosing (9–12) suggested that extending the standard monthly infusion to an interval of six weeks or longer could maintain therapeutic efficacy while reducing the risk of PML. However, the kinetics of the reconstitution of the cellular components of bone-marrow derived immune system in the CNS over the interval period between the natalizumab doses remained unclear.

In this study, we sampled the cerebrospinal fluid (CSF) in natalizumab-treated individuals and profiled its cellular component. By sampling CSF from individuals treated at different intervals, we investigated which cell populations are changing as the interval between doses is extended; this may provide insights to understanding CNS immune surveillance and that could guide the development of an optimal dosing interval.

## Results

**Figure 1A** outlines the design of our study based at Columbia University in New York (NY), with recruitment predominantly from the New York University MS Center and the Columbia MS Center, as well as referrals from other regional centers. The characteristics of the participants with relapsing-remitting MS (RRMS) based on the 2017 McDonald diagnostic criteria are outlined in **Supplemental Table 1**. Among the NY participants, three categories of persons with MS treated with natalizumab underwent CSF sampling: individuals treated with intravenous natalizumab 300 mg every (a) 4 weeks (n=7), (b) 6 weeks (n=11), or (c) 8 weeks (n=5). The participants had been treated for > 1 year on their respective regimens at the time of CSF sampling, and sampling was performed up to 3 days before the next scheduled dose of natalizumab. We also sampled 4 untreated individuals with RRMS who were sampled at the time of a diagnostic lumbar puncture to establish a reference CSF profile of MS. Additionally, we sampled 1 individual without MS but diagnosed with PML in the context of anti-rejection medication for organ transplantation (hence, unrelated to natalizumab) and 1 individual with RRMS treated with ocrelizumab with suspected PML (later ruled out by biopsy).

**Figure 1.**
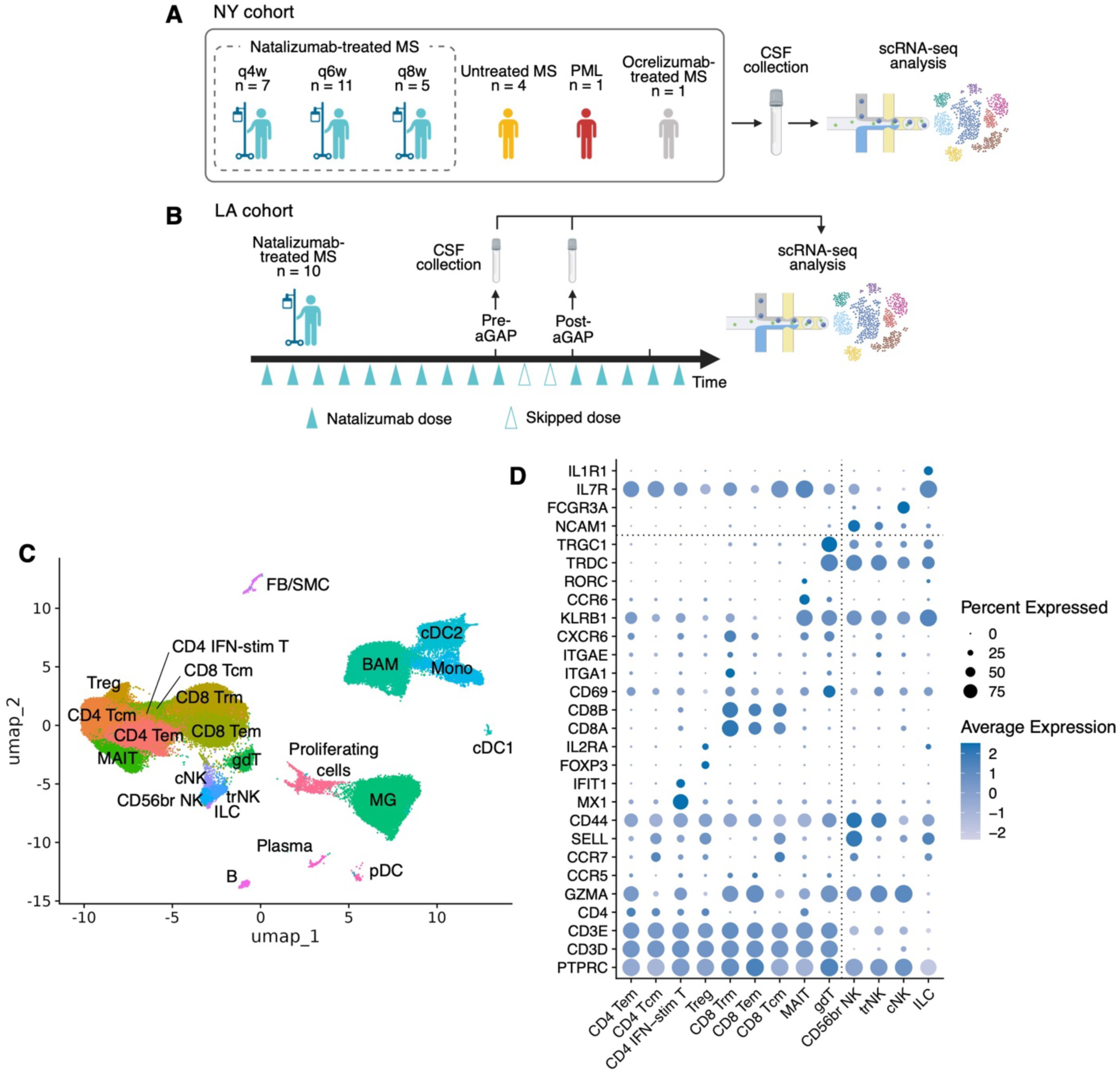
Single-cell transcriptome of CSF from natalizumab-treated MS patients. (**A**, **B**) Schema of this study. (**A**) Sample collection and analysis in the NY cohort. q4w, SID of 4 week interval; q6w, EID of 6 week interval; q8w, EID of 8 week interval. (**B**) Sample collection and analysis in the aGAP LA cohort. (**C**) UMAP dimensionality reduction of 99,102 cells in 46 CSF specimens from 39 patients. Cells are colored by their cell types. Tem, effector memory T cells; Tcm, central memory T cells; IFN-stim T, interferon-stimulated T cells; Treg, regulatory T cells; Trm, tissue-resident memory T cells; MAIT, mucosal-associated invariant T cells; gdT, γδ T cells; CD56br NK, CD56^bright^ NK cells; trNK, tissue-resident NK cells; cNK, conventional NK cells; ILC, innate lymphoid cells; B, B cells, Plasma, plasma cells; MG, microglia; BAM, border-associated macrophage; Mono, monocytes; cDC1, conventional type 1 dendritic cells; cDC2, conventional type 2 dendritic cells; pDC, plasmacytoid dendritic cells; FB, fibroblasts; SMC, smooth muscle cells; Prolif., proliferating cells. (**D**) Marker gene expression in clusters of T cells and NK cells. Markers of the other cell types are shown in **Supplemental** Figure 1.

We also assessed data collected in a parallel study performed at a private practice in Los Angeles (LA) (**Figure 1B**). In the LA cohort (n=10), the RRMS participants follow a different clinical protocol, receiving natalizumab every 4 weeks for 11 doses followed by an annual treatment-free gap of 12 weeks (two deferred infusions, effectively) before resuming every 4-week dosing (aGAP)(13). CSF was sampled twice: first before the last monthly dose and then 12 weeks later, at the end of the 12-week aGAP. In all cases, CSF cells were collected and processed for single-cell RNA sequencing within 4 hours of lumbar puncture (details in the **Methods**).

Following pre-processing and quality control, 46 samples from 39 participants yielded usable data, yielding a total of 99,102 transcriptomes from individual CSF cells that were considered in downstream analyses. On average, each sample yielded 2,154 transcriptomes after quality control. **Figure 1C** presents a Uniform Manifold Approximation and Projection (UMAP) dimensionality reduction where cells were clustered using the Louvain algorithm and annotated with the help of the annotateCSF software (14). In the data set as a whole, we identified the major expected classes of lymphoid, myeloid, and other cell types and subtypes that have been reported in the CSF (14–17). **Figure 1D** and **Supplemental Figure 1** show the expression levels of marker genes in cell types.

### Effect of natalizumab and reconstitution in the NY cohort

We first evaluated which cell populations differed in the context of natalizumab treatment. Specifically, we found differences in the proportions of eight cell subtypes when comparing samples from patients receiving any natalizumab dosage with profiles seen in untreated persons with MS sampled at the time of their diagnostic evaluation (**Figure 2**). Among T cell populations, we found significantly lower (FDR-adjusted p-value <0.05) proportions of central memory T cells (TCM)(CD4+ and CD8+), effector memory CD4+ T cells (TEM), and regulatory T cells (Tregs) in patients receiving natalizumab, along with higher proportions of tissue-resident memory CD8+ T cells (TRM). Among myeloid cells, we observed lower proportions of plasmacytoid dendritic cells (pDC) and higher proportions of microglia-like (MG-like) cells. Finally, there was a lower proportion of B cells as well as a suggestion that plasma cells are also lower (nominal p<0.05). Because our analysis was based on proportions rather than absolute cell numbers, the higher proportion of certain tissue-resident cell types (cells that may have been homing in the CNS for a long time, i.e., TRM and MG-like cells) may simply reflect the reduction of other cell populations rather than a true increased migration of these cells into the CSF.

**Figure 2.**
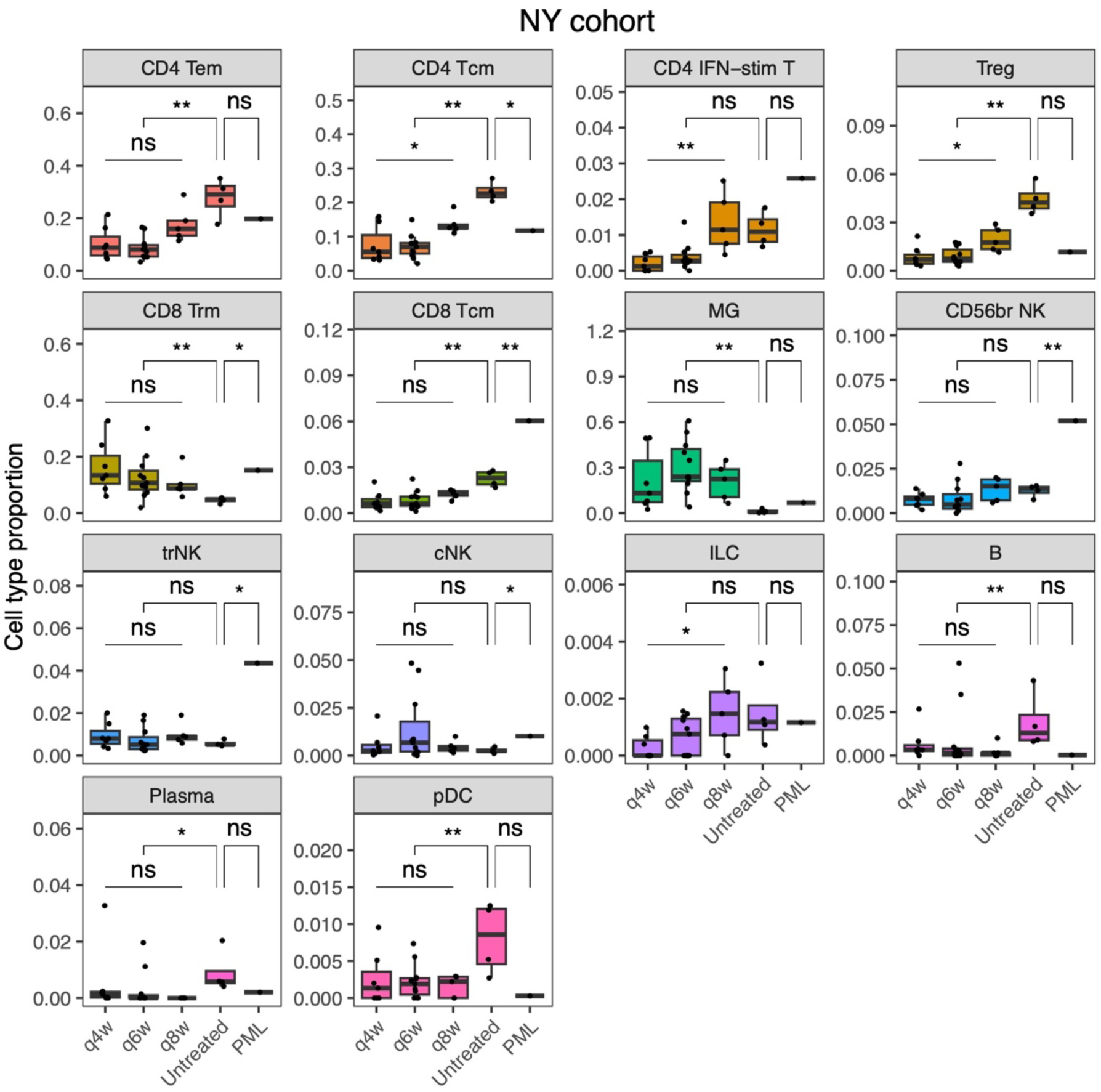
Cell type proportions in CSF of the NY cohort. Dots represent CSF specimens. Elements of the boxes show the following statistics: center line, median; box limits, upper and lower quartiles; whiskers, the most extreme data point which is no more than 1.5 times the interquartile range from the box. The *p*-values were computed with a linear model adjusted for age and sex. Untreated, untreated MS; **, FDR < 0.05; *, nominal *p*-value < 0.05; ns, non-significant. Only cell types that have at least one statistically significant result are shown here. The other cell types are shown in **Supplemental** Figure 2.

Having characterized the overall composition of CSF cell populations in natalizumab-treated participants, we then assessed whether there was a linear trend for a change in the proportion of each cell subtype over time since the last infusion using the three groups of treated participants: 4 week, 6 week, and 8 week interval dosing. We use this approach to investigate the trends of immune reconstitution following a natalizumabinfusion, in relapse-free individuals treated for >1 year with natalizumab. **Figure 2** illustrates that only one cell population, CD4+ T cells with an interferon response signature (CD4+IFN+ T), showed a significant increase in proportion (FDR<0.05), with a rise that was most pronounced between the 6 week and 8 week timepoints. By 8 weeks, the average proportion of this T cell subtype had attained the proportions seen in untreated individuals. Further, three cell subtypes displayed suggestive evidence of change over the extended interval (nominal p<0.05): innate lymphoid cells (ILC) are seen to gradually increase in proportion to untreated MS levels by the 8 week time point, while Treg and CD4+ TCM trend toward but did not reach the untreated levels.

### Observation of the CSF cellular constituents across a treatment gap

We then considered data from the LA aGAP cohort in which individuals were sampled before their last monthly infusion from a series of 11 infusions of the standard interval (4 weeks) for natalizumab dosing as well as at the end of the aGAP of 12 weeks. Here, we saw some heterogeneity in the kinetics of the reconstitution among the 6 participants who underwent 2 lumbar punctures (before the last pre-gap infusion and at the end of the aGAP). **(Figure 3**). Evaluating first the CD4+IFN+ T cell population that was significant in the prior analysis (**Figure 2**), we saw that 4 of the 6 participants had an increase in the proportion of this cell type, consistent with the prior result, but the trend was not statistically significant, suggesting that a larger sample size might be needed to explore this further. In these individuals with longitudinal data, there was a significant (FDR<0.05) decrease in the proportion of CD8+ TRM and increase in the proportion of CD56^bright^ NK cells (**Figure 3**). As in the NY sample collection, the ILC, CD4+ TCM, and Treg populations had a suggestive level of evidence (nominal p<0.05) that their proportions were increasing over the longer interval. While these results did not meet our threshold of significance, the consistency of the effect in these two very different analyses (NY cross-sectional sample collection and LA longitudinal data) is notable.

**Figure 3.**
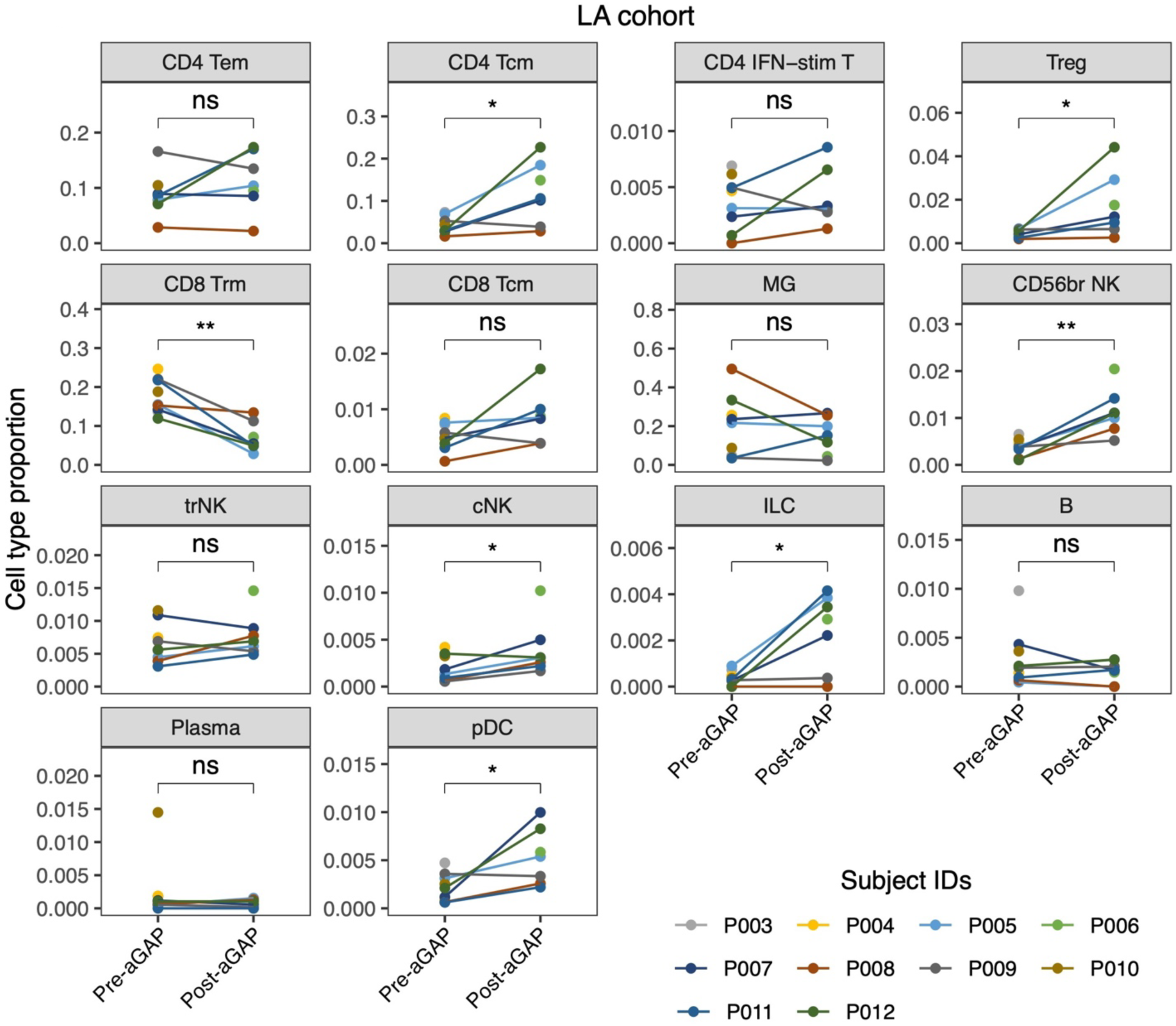
Cell type proportions in CSF of the LA cohort. Dots represent CSF specimens. Pre- and post-12 week aGAP CSF specimens of the same patient are connected with a line. Only pre-aGAP or post-aGAP specimens were available in some patients. **, FDR < 0.05; *, nominal *p*-value < 0.05; ns, non-significant. The same cell types with Figure 2 are shown here. The remaining cell types are shown in **Supplemental** Figure 3.

### Relation to PML

Over the course of the recruitment for the NY cohort, we were able to profile 1 individual with PML in the context of anti-rejection medication for organ transplantation. JC viral load in CSF was low but detectable the week prior to the CSF sample collection, and the participant’s infection was ultimately controlled with reduction of his immunosuppressive treatment. When comparing the profile of this individual with that of untreated MS participants using a linear model adjusted for age and sex (**Figure 2**), we found that CD8+ TCM cells showed an increased proportion, along with CD56^bright^ NK cells. There was also suggestive evidence for an increase of CD8+ TRM, tissue-resident NK (trNK), and conventional NK (cNK) cells; there was also a reduction in the proportion of CD4+ TCM. In relating these observations to our earlier results, the trNK population stood out as it was not altered in any of the preceding analyses, and this change seems specific to the context of PML in this case with burgeoning viral control. The CD56^bright^ NK cells were also increased in the analysis of the longitudinal samples following an annual12-week natalizumab infusion gap (**Figure 3**). Both trNK and CD56^bright^ NK cell types were therefore associated with PML viral control and reconstitution of the CSF cell populations following natalizumab, making them prime candidates for further evaluation in the context of titrating the interval of natalizumab dosing to reduce the likelihood of PML.

### Changes in gene expression during immune reconstitution

To complement the evaluation of cell type proportions, we also performed differential gene expression analysis within each of our cell types. **Figure 4A** reports the summary of our analyses across the cell subtypes: the largest difference in gene expression is seen between untreated and natalizumab-treated RRMS participants. We compared EID and SID regimens in the NY cohort, and we compared these results to the results of a longitudinal analysis pre- and post- aGAP in the LA samples. We found that the border-associated macrophage (BAM) population had the largest number of changes in gene expression relative to other cell types (**Figure 4B**) in both analyses. However, the list of differentially expressed genes from these two analyses of BAMs were largely different (**Supplemental Figure 4A**), suggesting that there may be differences that emerge between 8 and 12 weeks post-natalizumab. There were only two shared differentially expressed genes (DEGs) between the cohorts, which were found in CD56^bright^ NK cells (**Supplemental Figure 4B**). Further, it is notable that the smaller LA collection of samples uncovered many more differences in gene expression in multiple cell types, and the same pathways related to viruses and interferon response were differentially expressed in multiple cell types (**Figure 4C**). The longitudinal analysis is less sensitive to inter-individual heterogeneity, which may, in part, explain the larger number of genes detected, but it may also reflect the fact that the longer interval of time since the last infusion of natalizumab has altered the CNS context and led to a relative upregulation of anti-viral pathways.

**Figure 4.**
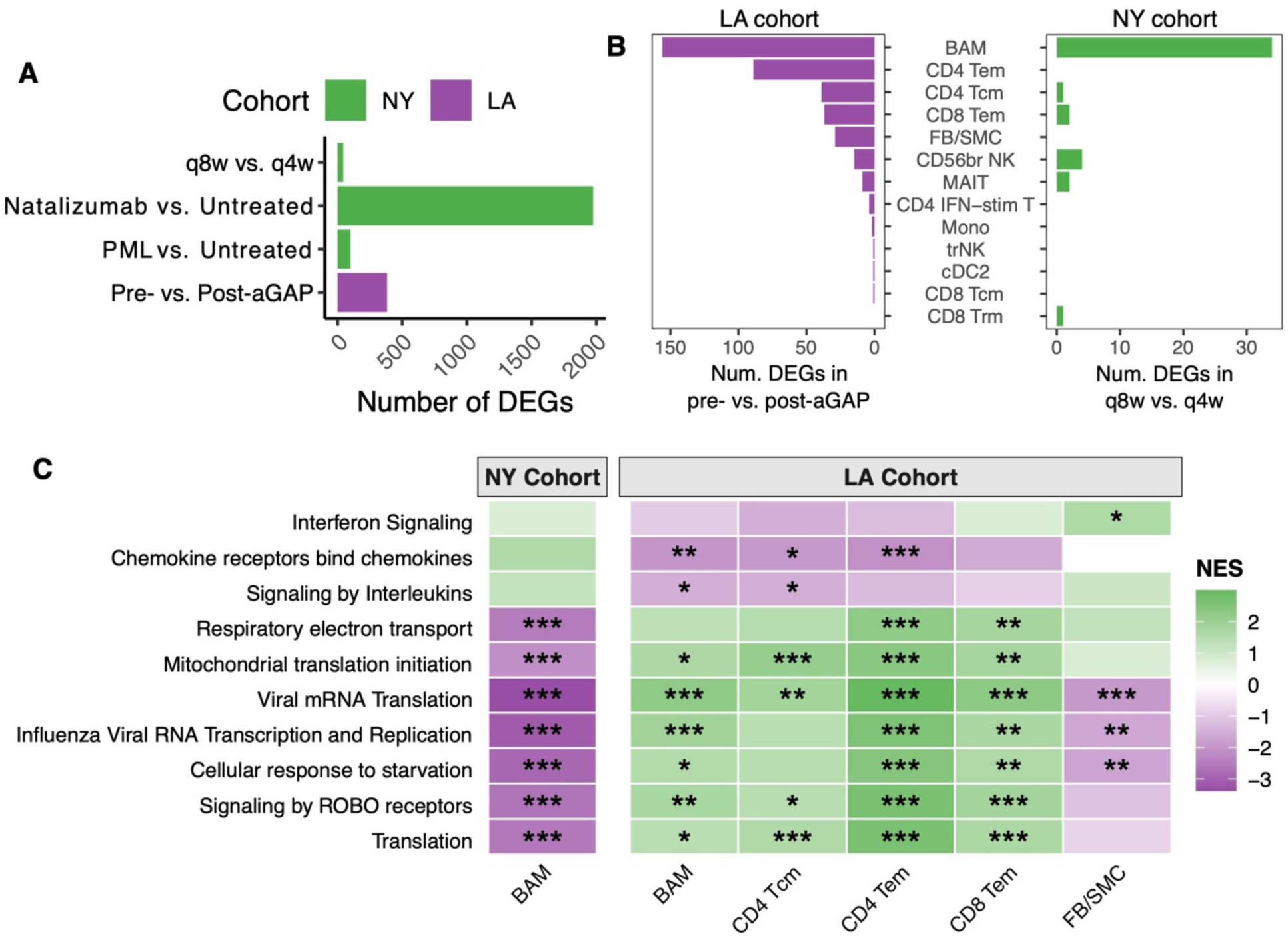
Cell type-level differences of gene expression between MS treatment. (**A**) The total number of differentially expressed genes (DEGs) between conditions. Cell type-level DEGs were computed, and their numbers were aggregated over all cell types. (**B**) The number of cell type-level DEGs between EID and SID in the two cohorts. In the LA cohort, pre-12 week gap is SID, and post-12 week gap is aGAP. In the NY cohort, we used q8w as EID here because there were few DEGs between q6w and q4w. (**C**) Pathway enrichment of cell type-level DEGs. Gene set enrichment analysis was applied to ranked gene lists of cell type-specific differential expression using the Reactome pathway database. Normalized enrichment score (NES) is positive when the pathway is upregulated in EID of the NY cohort or post-aGAP samples of the LA cohort. ***, q-value < 0.001; **, q-value < 0.01; *, q < 0.05.

### In vitro endothelial re-adhesion of CD4+ TCM in EID patients

Previous studies have shown the increased desaturation of natalizumab receptors in peripheral lymphocytes in EID compared to SID (18), which is believed to increase the re-adhesion of lymphocytes to the CNS endothelium in EID. To further investigate the relative cell-type-specificity of the re-adhesion, we performed perpetual physiological flow adhesion assays to primary human CNS endothelium with previously cryopreserved PBMCs from SID and EID patients (**Figure 5A**). T cells from EID patients showed lower levels of natalizumab saturation compared to SID patients (**Figure 5B**), indicating an increased potential of these cells to use VLA-4 to re-adhere to the endothelium. Flow cytometric analysis of the adhered cells revealed that the CD4/CD8 ratio was higher in EID patients (**Figure 5C**). Consistent with the single-cell RNA-seq results from the CSF, further analysis of the CD4+ T cells showed a higher proportion of adhered CD4+ TCM in EID compared to SID patients (**Figure 5D**), whereas the proportion of CD4+ TEM did not differ between both patient groups (**Figure 5E**). This indicates that some CD4+ TCM may indeed show an increased potential to re-adhere to CNS endothelium and re-enter the CNS in natalizumab-treated RRMS patients on an EID.

**Figure 5.**
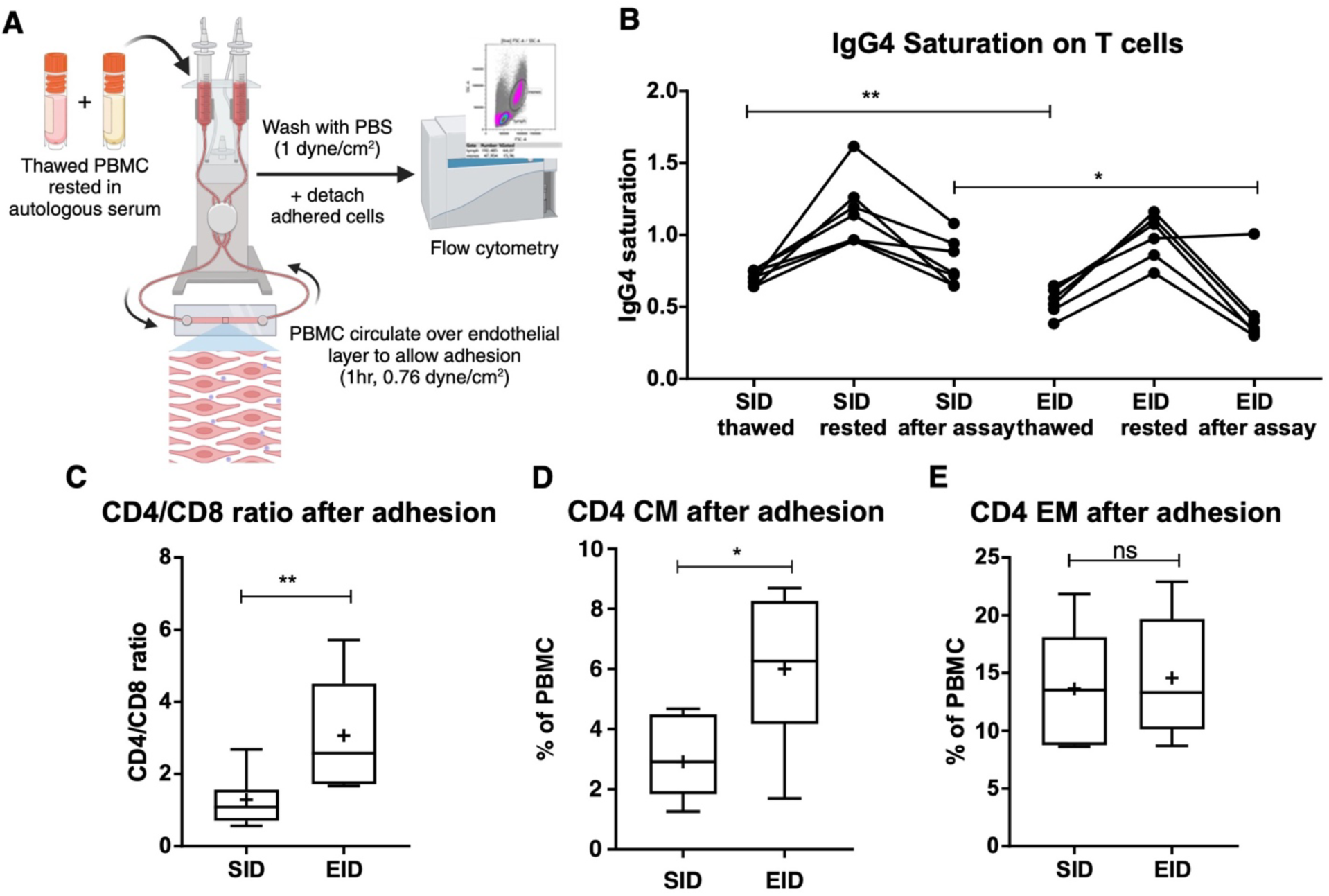
In vitro endothelial re-adhesion of CD4+ TCM in EID and SID patients. (**A**) Schematic of the assay procedure with thawed PBMC from EID versus SID patients rested in autologous serum, then circulated over a layer of primary human endothelium for 1h at 0.76 dyne/cm2, after which the adherent cells are harvested and analyzed using flow cytometry. (**B**) Shown is the level of IgG4 saturation in relation to sample aliquots spiked with natalizumab to delineate 100% saturation. “Thawed” are saturation values directly after thawing the PBMC, “rested” are saturation values after resting the PBMC overnight at 37°C in autologous patient serum, “after assay” are saturation values after harvesting the cells. (**C**) Shown is the CD4/CD8 ratio after the adhesion assay in SID versus EID patients. (**D**) Shown is the percentage of CD4+ TCM of harvested PBMC after the adhesion assay in SID versus EID patients. (**E**) Shown is the percentage of CD4+ TEM cells of harvested PBMC after the adhesion assay in SID versus EID patients. *, p-value < 0.05; **, p-value < 0.01; ns, non-significant. Boxes indicate 25% and 75% percentiles, the line indicates the median, + indicates the mean, and whiskers indicate the minimum/maximum values.

## Discussion

In this study, we investigated the characteristics of immune reconstitution in the CSF following natalizumab immunotherapy. Natalizumab markedly alters the cellular constituents of the CSF compared with untreated MS. The most dramatic changes are the reductions in the proportions of CD4+ and CD8+ TCM, CD4+ TEM, Tregs, B cells and pDCs. This reduction is aligned with the known mechanism of natalizumab, i.e. blocking trafficking of leukocytes from the peripheral circulation into the CNS. Our observations align with prior work showing that CD4⁺ T-cell help maintains effective brain-resident CD8⁺ responses and that dysfunction in either subset undermines control of JC virus, predisposing to PML(19, 20). Conversely, MG-like cells and TRM CD8+ T cells are proportionally increased in natalizumab-treated samples, presumably reflecting their resident status rather than trafficking into the CNS(14).

As the treatment interval is extended, there is gradual recovery of some cellular populations including the CD4+ TCM, CD4+IFN+ T, Treg, and ILC, supporting a time dependent immune reconstitution. Comparison between the EID immune reconstitution pattern and the PML case immune profile suggests that the demonstrated PML reduction in EID (13) may be related to the return of the CD8+ TCM. The PML patient who was sampled had a minimal CSF JC viral load at the time of lumbar puncture and was essentially controlling the infection (and subsequently recovered from it). In addition to the difference in CD8+ TCM, this participant had a notable relative elevation in CD56^bright^ NK cells, which were also interestingly elevated after the annual 12-week gap in natalizumab infusion that was implemented in the LA cohort, suggesting an additional role for CD56^bright^ NK cells in viral control.

The adhesion assay validates the *ex vivo* CSF finding *in vitro* by showing relative potential for CD4+ TCM from EID or SID patients to re-enter the CNS. It is also consistent with the hypothesis that EID is more compatible with enhanced re-adhesion capacity and presumably a greater likelihood of extravasation into the perivascular and parenchymal compartments(21). This is supported by the elevated CD4:CD8 ratio on flow cytometry in the CSF of EID patients and mirrors the single cell data from the NY and LA cohorts.

Taken together, our work suggests that this earlier return of some CD4+ T cells may contribute to enhanced antiviral competence while other lymphocytic subtypes remain relatively excluded, potentially diminishing the risk of PML while preserving the efficacy of the treatment. In addition to lymphocyte dynamics, it is increasingly recognized that border-associated macrophages (BAMs) contribute substantially to immune regulation at CNS interfaces. As highlighted by Mundt and colleagues, BAMs function as critical gatekeepers that sense peripheral immune activity, modulate leukocyte entry, and maintain vascular and meningeal homeostasis(22).

This study highlights the waning action of natalizumab with advancing time. These immune reconstitution patterns provide insights into the mechanisms of viral control, while the cells that are persistently excluded may be more responsible for the inflammatory activity of relapsing MS given the successful suppression of relapse with EID regimen (14) and the aGAP approach (12).

## Methods

The institutional review board of Columbia University Irving Medical Center approved the study protocol (IRB number: AAAR9480). Informed consent was obtained from all participants prior to their inclusion. Participant confidentiality and data privacy were maintained in accordance with institutional standards and the guidelines of the Federal Office for Human Research Protections.

CSF samples were obtained with lumbar punctures and centrifuged to collect the cell pellet. Single-cell RNA-seq libraries were prepared from resuspended cells using the Chromium Next GEM Single Cell 3’ v3. Details are described in **Supplementary Methods**.

## Data availability

The count matrix of single-cell RNA-seq generated by CellRanger can be obtained from the public repository Figshare with the following link: https://doi.org/10.6084/m9.figshare.30854225.

## Acknowledgements

This study was supported by research funds from Biogen. The investigators acknowledge the generosity of the individuals who participated in this study.

## References

1. Polman CH, et al. A randomized, placebo-controlled trial of natalizumab for relapsing multiple sclerosis. N Engl J Med. 2006;354(9):899–910.

2. McEntire CRS, et al. Characteristics of Progressive Multifocal Leukoencephalopathy Associated With Sarcoidosis Without Therapeutic Immune Suppression. JAMA Neurol. 2023;80(6):624–633.

3. Aggarwal D, et al. Progressive multifocal leukoencephalopathy in idiopathic CD4(+) lymphocytopenia: A case report and review of literature. Neuropathol Off J Jpn Soc Neuropathol. 2019;39(6):467–473.

4. Berger JR. Progressive multifocal leukoencephalopathy in acquired immunodeficiency syndrome: explaining the high incidence and disproportionate frequency of the illness relative to other immunosuppressive conditions. J Neurovirol. 2003;9 Suppl 1:38–41.

5. Khoury SJ, et al. Changes in activated T cells in the blood correlate with disease activity in multiple sclerosis. Arch Neurol. 2000;57(8):1183–1189.

6. Bozic C, et al. Anti-John Cunnigham virus antibody prevalence in multiple sclerosis patients: baseline results of STRATIFY-1. Ann Neurol. 2011;70(5):742–750.

7. Plavina T, et al. Anti-JC virus antibody levels in serum or plasma further define risk of natalizumab-associated progressive multifocal leukoencephalopathy. Ann Neurol. 2014;76(6):802–812.

8. Tan IL, et al. Immune reconstitution inflammatory syndrome in natalizumab-associated PML. Neurology. 2011;77(11):1061–1067.

9. Foley JF, et al. Comparison of switching to 6-week dosing of natalizumab versus continuing with 4-week dosing in patients with relapsing-remitting multiple sclerosis (NOVA): a randomised, controlled, open-label, phase 3b trial. Lancet Neurol. 2022;21(7):608–619.

10. Ryerson LZ, et al. Risk of natalizumab-associated PML in patients with MS is reduced with extended interval dosing. Neurology. 2019;93(15):e1452–e1462.

11. Ruggieri S, et al. Treatment modifiers across different regimens of natalizumab treatment in MS: An Italian real-world experience. Neurother J Am Soc Exp Neurother. 2024;21(3):e00338.

12. Berkovich R, et al. CD4 cell response to interval therapy with natalizumab. Ann Clin Transl Neurol. 2015;2(5):570–574.

13. Berkovich R, et al. Annual 12-Week Dosing Gap of Natalizumab: Clinical Efficacy, Blood Biomarkers, and CSF Cell Composition. Ann Clin Transl Neurol. [published online ahead of print: September 23, 2025]. 10.1002/acn3.70207.

14. Ostkamp P, et al. A single-cell analysis framework allows for characterization of CSF leukocytes and their tissue of origin in multiple sclerosis. Sci Transl Med. 2022;14(673):eadc9778.

15. Touil H, et al. A structured evaluation of cryopreservation in generating single-cell transcriptomes from cerebrospinal fluid. Cell Rep Methods. 2023;3(7):100533.

16. Schafflick D, et al. Integrated single cell analysis of blood and cerebrospinal fluid leukocytes in multiple sclerosis. Nat Commun. 2020;11(1):247.

17. Farhadian SF, et al. Single-cell RNA sequencing reveals microglia-like cells in cerebrospinal fluid during virologically suppressed HIV. JCI Insight. 2018;3(18):121718.

18. Zhovtis Ryerson L, et al. Pharmacodynamics of natalizumab extended interval dosing in MS. Neurol Neuroimmunol Neuroinflammation. 2020;7(2). 10.1212/NXI.0000000000000672.

19. Mockus TE, et al. CD4 T cells control development and maintenance of brain-resident CD8 T cells during polyomavirus infection. PLoS Pathog. 2018;14(10):e1007365.

20. Jilek S, et al. Immune responses to JC virus in patients with multiple sclerosis treated with natalizumab: a cross-sectional and longitudinal study. Lancet Neurol. 2010;9(3):264–272.

21. Herich S, et al. Human CCR5high effector memory cells perform CNS parenchymal immune surveillance via GZMK-mediated transendothelial diapedesis. Brain J Neurol. 2019;142(11):3411–3427.

22. Mundt S, Greter M, Becher B. The CNS mononuclear phagocyte system in health and disease. Neuron. 2022;110(21):3497–3512.

