## Supplemental Data 1 for "Cerebrospinal fluid cellular dynamics following modified interval dosing natalizumab therapy in multiple sclerosis"

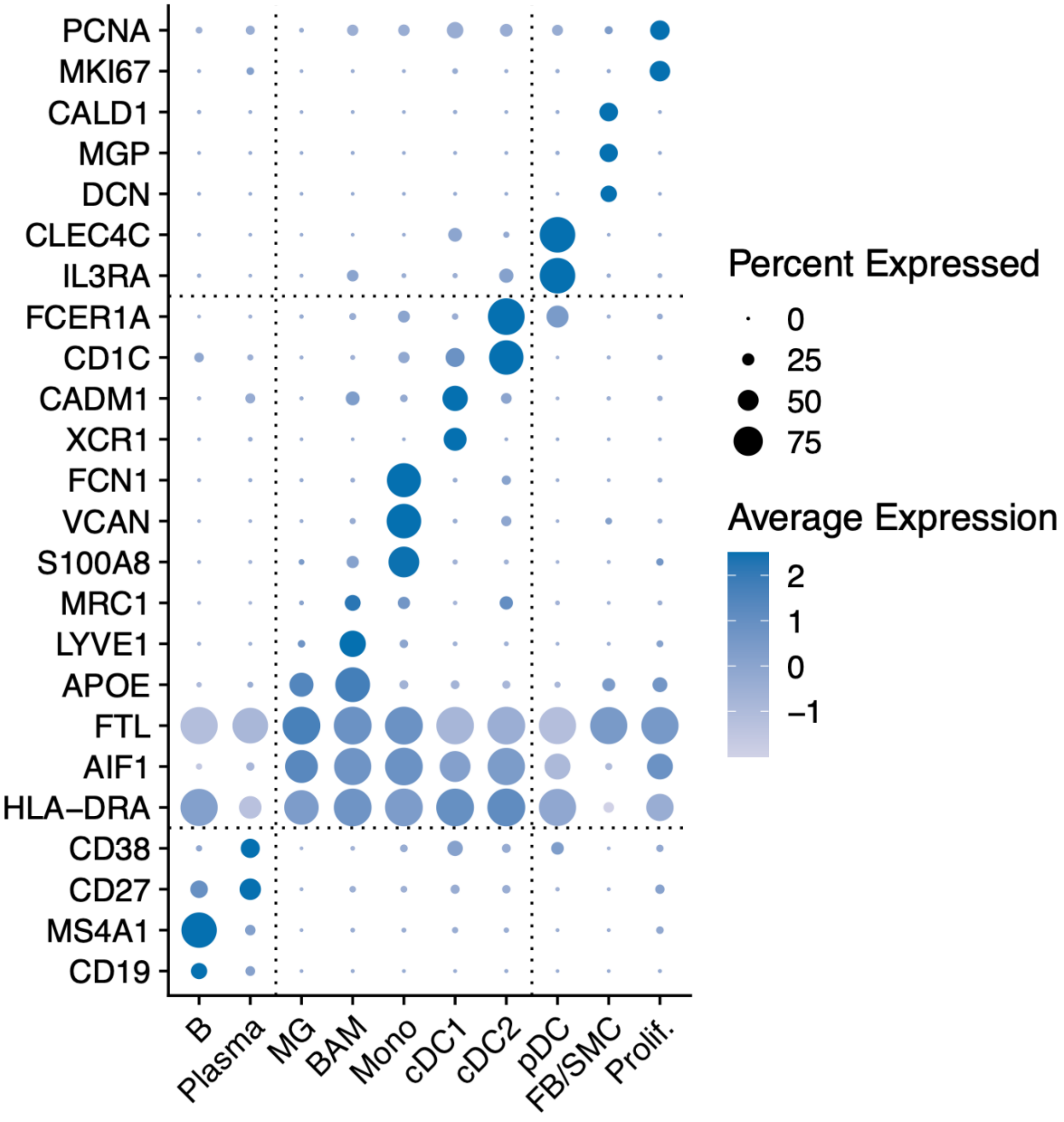


**Supplemental Figure 1**. **Marker gene expression in clusters of B cells, myeloid cells, and miscellaneous cells.**

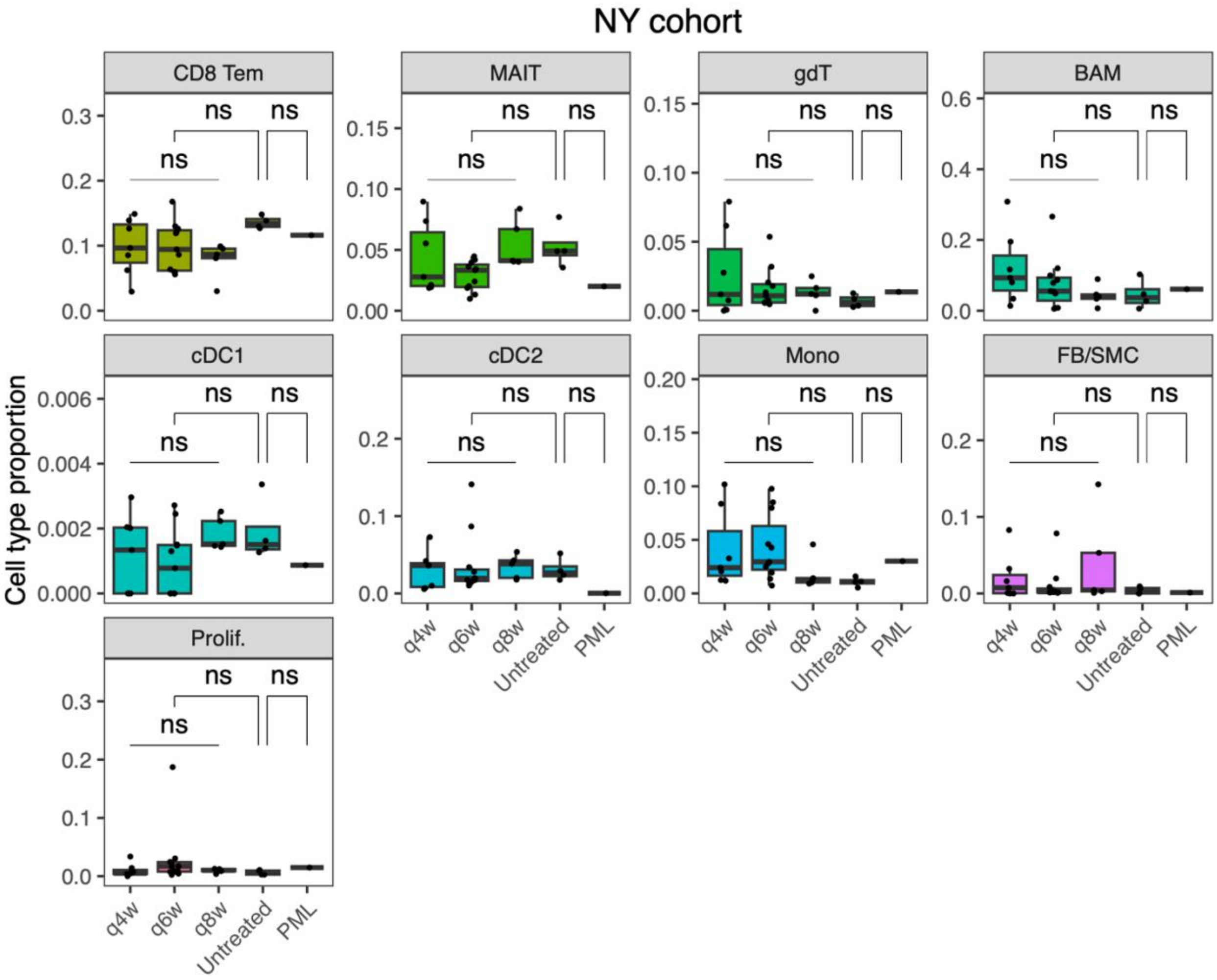


**Supplemental Figure 2**. **Non-significant cell type proportions of the NY cohort**. Dots represent CSF specimens. Elements of the boxes show the following statistics: center line, median; box limits, upper and lower quartiles; whiskers, the most extreme data point which is no more than 1.5 times the interquartile range from the box. The *p*-values were computed with a linear model adjusted for age and sex. Untreated, untreated MS; ns, non-significant.

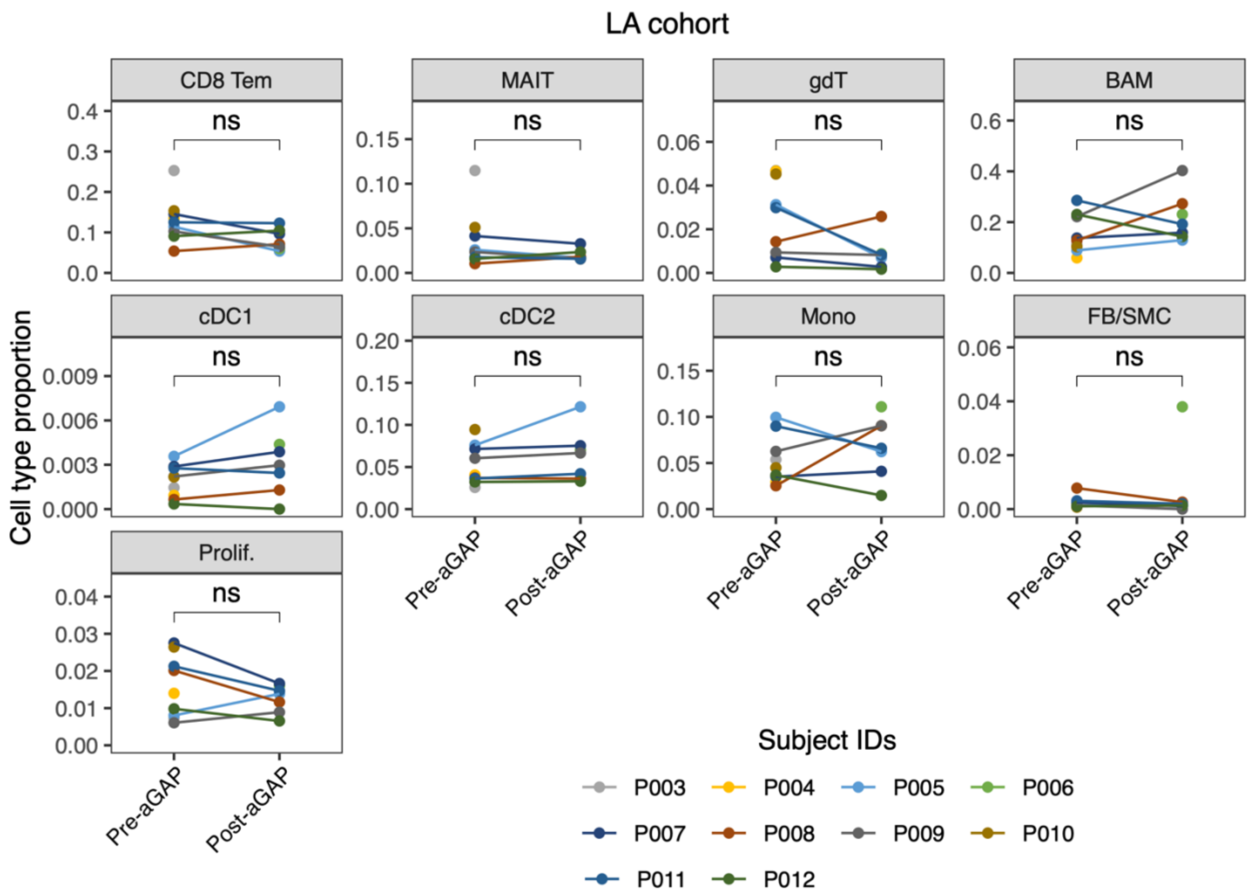


**Supplemental Figure 3**. **Non-significant cell type proportions of the LA cohort**. Dots represent CSF specimens. Pre- and post-12 week aGAP CSF specimens of the same patient are connected with a line. Only pre-aGAP or post-aGAP specimens were available in some patients. ns, non-significant.

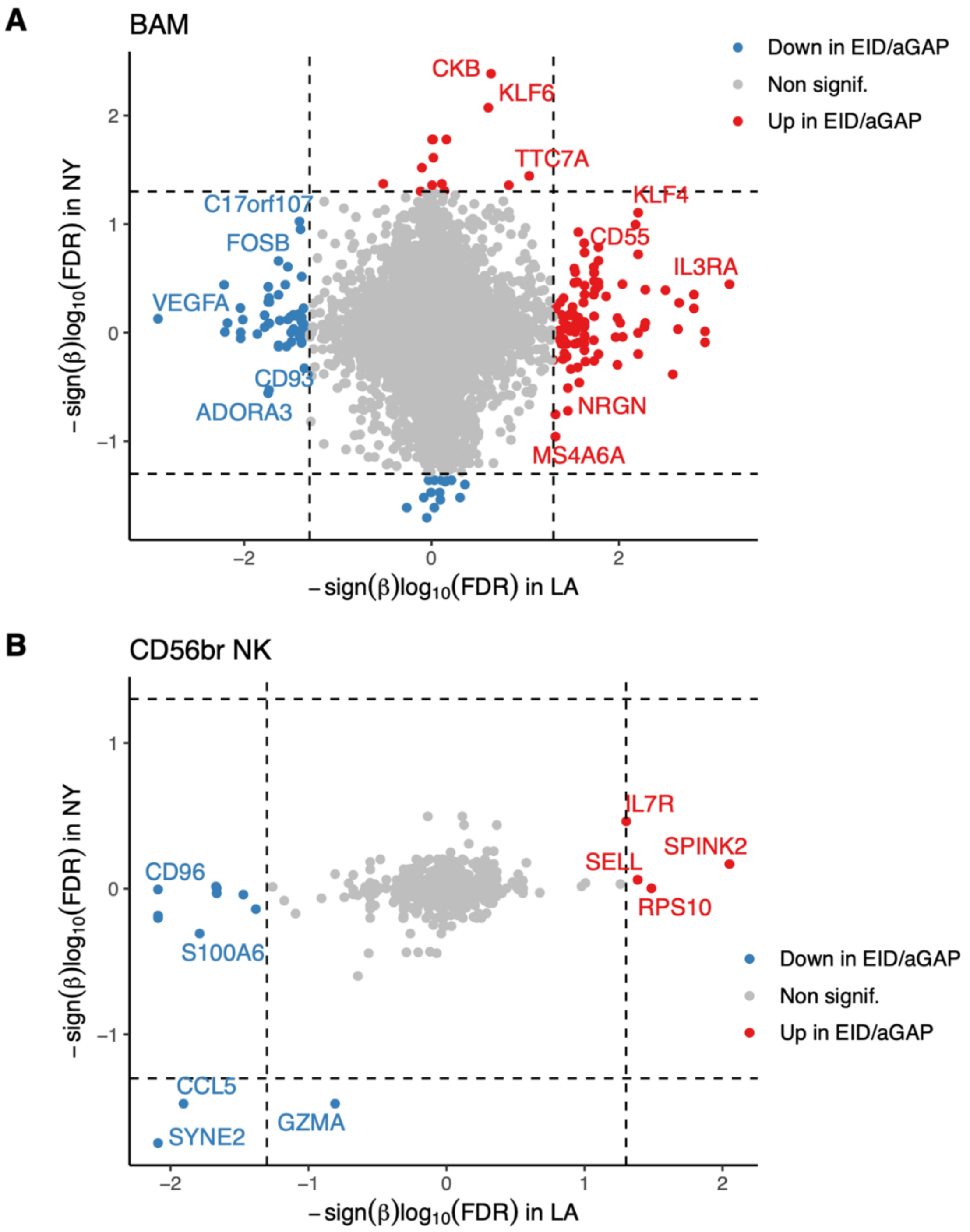


**Supplemental Figure 4**. **Limited overlap of DEGs between the NY and LA cohorts.** Cell type-specific DEGs between EID (including post-aGAP) and SID were compared across the two cohorts in terms of their effect directions and statistical significance. (**A**) DEGs specific to BAM. (**B**) DEGs specific to CD56$^{bright}$ NK cells. $\beta$ is a log-fold change of gene expression in EID relative to SID. Red, genes that have higher expression in EID than SID; blue, genes lower in EID than SID; gray, non-significant genes. Dotted lines represent FDR = 0.05.
