## Supplementary Methods for "Cerebrospinal fluid cellular dynamics following modified interval dosing natalizumab therapy in multiple sclerosis"

Sex as a biological variable: We observed no significant interaction between sex and natalizumab dosing intervals in all cell types of both cohorts.

**Lumbar puncture and CSF sample processing**

Lumbar punctures were performed using atraumatic Sprotte spinal needle. CSF samples were then immediately transported to the laboratory in an ice bucket to maintain a stable temperature of 0ºC to 4ºC. CSF was centrifuged at 300g for 10 minutes at 4ºC to collect the cell pellet which was resuspended in 70µL of autologous CSF prior to loading on the 10x Genomics Chromium platform. All samples used in this study were processed fresh immediately after collection and were loaded on the Chromium platform within 4 hours of CSF extraction. Single-cell suspensions were loaded onto the 10x Genomics Chromium Controller for barcoding and library preparation using the Chromium Next GEM Single Cell 3’ v3. Processing was performed according to manufacturer’s instructions. Sequencing was performed using Illumina NextSeq 500 with 26-8-0-57 read setup.

**Single-cell RNA-seq data processing**

Sequencing data from all samples were processed using a single pipeline. Data from each sample was processed separately before integration. Cell Ranger v3.1 was used for aligning the sequenced reads to reference 3.1.0 (Ensembl 93) transcriptome, unique molecular identifier (UMI) quantification, and cell calling. Doublets were identified using Scrublet. The expected percentage of doublets/multiplets was estimated according to the 10x Genomics Single Cell User Guides proportional to the number of recovered cells. The doublet score threshold was set at the minimum between the two peaks of the bimodal simulated doublet score histogram. The rest of the pre-processing was performed using Seurat v4.0.1. Data from each sample underwent normalization and principal component analysis (PCA) using default parameters. The first 20 PCs were used to perform an initial clustering in order to identify the red blood cell (RBC) clusters (i.e., clusters with high expression of hemoglobin genes). RBC clusters were then removed from further analysis along with the Scrublet-identified doublets and additional low-quality cells (i.e., cells with >25% mitochondrial transcripts, <100 unique identified genes or >20 UMI/gene ratio). Ambient RNA contamination removal was performed using SoupX.

Gene expression levels were integrated across libraries using the IntegrateLayers function of Seurat with the Harmony method. Low-resolution clustering of cells was performed with the FindClusters function with resolution = 0.2. We ran the annotateCSF software (v1.1a) on this dataset and annotated clusters with major cell types with the help of annotateCSF lablels. To further subclassify T cells, the clusters of T cells were extracted, and FindClusters was applied with resolution = 0.4. Likewise, the clusters of myeloid cells and NK cells were subclustered with resolution = 0.1 and 0.2, respectively.

**Differential gene expression analysis**

Differential gene expression analysis was performed with R package limma (v3.62.1). For each cell type, a cell type-specific pseudobulk count matrix was constructed by aggregating UMI counts from the cell type per library. Low-expression genes were excluded with the filterByExpr function with default parameters, and library size was adjusted with the trimmed-mean-of-M-values normalization. The mean-variance relationship was modeled with the voom function, and p-values were computed with lmFit and eBayes. The paired nature of the LA samples was handled by incorporating a blocking variable and the correlation of duplicates to the lmFit function. Pathway enrichment analysis of differentially expressed genes was performed with the R package clusterProfiler (v4.14.6).

**Adhesion assay and IgG4 saturation staining**

Human brain microvascular endothelial cells (HBMECs, PeloBiotech) were cultivated in 0.4mm µ-slide I channel slides (ibidi) under continuous, unidirectional flow (shear stress: 0.5 – 2.0 dyne/cm^2^) in endothelial growth medium (PeloBiotech) using a fluidic pump system (ibidi) to generate HBMEC monolayers. After 48-72 hours, tumor necrosis factor-α (R&D Systems, final concentration 100 U/ml) was added and HBMECs were cultivated for another 18-20 h under continuous, unidirectional flow (2 dyne/cm^2^). PBMCs and 500 – 1000 µL serum from SID (n = 7) and EID (n = 6) natalizumab-treated RRMS patients were thawed and the PBMC were rested in autologous serum overnight at 37 °C to allow re-saturation with individual natalizumab concentration. The mean dosing interval over the last six months was 29.5 days in the SID group and 43.6 days in the EID group. Approximately 5 x 10^6^ PBMCs per patient were added to a fluidic unit with a HBMEC monolayer. A schematic overview of the assay is given in **Figure 5A**. The PBMCs circulated over the endothelial cells for one hour in a unidirectional flow (0.76 dyne/cm^2^). Subsequently, the slides were washed with phosphate-buffered saline (PBS) in a unidirectional flow (1.0 dyne/cm^2^) for 8 minutes to remove non-specifically adhered PBMCs. Afterwards, the channel bottoms were cut out of the slides and incubated with trypsin/EDTA (Sigma-Aldrich) on a test tube shaker at 37°C to detach HBMECs and adhered PBMCs. The reaction was stopped with RPMI 1640 supplemented with 10 % FBS and the cells were stained with the following antibodies: CD3-PE/Dazzle™594, CD4-BV605™, CD19-PerCP, CD45RA-PE/Cy7, CD197-APC/Cy7 (BioLegend), CD8-APC/AF700, CD14-PC5.5 (Beckman Coulter). The cells were analyzed by flow cytometry using Kaluza software (Beckman Coulter).
Additionally, the IgG4 saturation with natalizumab on T cells was determined immediately after thawing, post overnight resting in autologous serum and following the one-hour adhesion assay. For this, PBMCs were resuspended in PBS and incubated for 30 min at 4 °C with (spiked) or without (naïve) natalizumab (1:2000, Biogen). Afterwards, the cells were stained with the following antibodies: CD3-PE/Dazzle™594 (Biolegend), IgG4-PE (Southern Biotech) and applied to flow cytometry. The IgG4 saturation of T cells was calculated by dividing the percentage of IgG4+ T cells in the naïve sample by that percentage in the spiked sample.

**Statistical analysis**

In the NY cohort, a linear model of the square root of cell type proportion was fitted with MS treatments as an independent variable and age and sex as covariates. To examine a trend associated with dosing intervals, the intervals were treated as numerical variables (4 week, 6 week, and 8 week). In the LA cohort, a linear mixed effect model was fitted where the square root of cell type proportion is a dependent variable, sample collection time (Pre- and Post-aGAP) is an independent variable, and age and sex as covariates. Because of the longitudinal nature of the LA cohort, a random intercept of patient IDs was added to the model using R package nlme (v3.1-166). Multiple hypothesis correction was performed by applying the Benjamini-Hochberg method to nominal p-values.
